# Transmission of SARS-CoV-2 associated with cruise ship travel: protocol for a systematic review (Version 1)

**DOI:** 10.1101/2021.10.11.21264724

**Authors:** Elena C Rosca, Carl Heneghan, Elizabeth A Spencer, Jon Brassey, Annette Plüddemann, Igho J Onakpoya, David H Evans, John M Conly, Tom Jefferson

## Abstract

**Background:** Maritime and river travel, including cruise ships, have been implicated with spreading viruses through infected passengers and crew. Given the novelty of the SARS-CoV-2 infection, early cruise ship travel transmission models of spread are based on what is known of the dynamics of other respiratory viral infections. Our objective is to provide a rapid summary and evaluation of relevant data on SARS-CoV-2 transmission aboard cruise ships, report policy implications, and highlight research gaps requiring attention.

**Methods:** We will search LitCovid, medRxiv, Google Scholar, and the WHO Covid-19 database using COVID-19, SARS-CoV-2, transmission, and cruise ship appropriate synonyms. We will also search the reference lists of included studies for additional relevant studies. We will include studies reporting onboard SARS-CoV-2 transmission from passengers and/or crew to passengers and/or crew. We will consider any potential transmission mode. We will assess study quality based on five criteria and report important findings. The outcome will consist of the onboard cruise ships’ transmission of SARS-CoV-2. We will provide a narrative summary of the data and report the outcomes, including quantitative estimates where feasible and relevant. Where possible, compatible datasets may be pooled for meta-analysis.

**Expected results:** We will present the evidence in three distinct packages: study description, methodological quality assessment and data extracted. We will summarize the evidence and will draw conclusions as to the quality of the evidence.

## Background

Coronavirus disease 2019 (COVID-19) is a novel condition, distinct from other diseases caused by coronaviruses, such as Severe Acute Respiratory Syndrome (SARS) and Middle East Respiratory Syndrome (MERS). The SARS-CoV-2 virus spreads rapidly, and the modes of transmission are not yet completely understood.

National and international organizations, including the WHO, are working to coordinate the development of prevention, control, and management measures on many fronts. The overarching aim is to control the SARS-CoV-2 pandemic by reducing transmission of the virus and preventing associated illness and death [WHO 2020]. Nevertheless, the transmission of the SARS-CoV-2 virus and the disease it causes are not entirely understood, and public health measures for restricting transmission are based on the best available information.

Maritime and river travel, including cruise ships, have been implicated with spreading viruses through infected passengers and crew. Firstly, cruise ships, with their closed or partially enclosed environments, may facilitate virus transmission because passengers and crew are in close proximity for extended periods [Millman 2015]. The shipboard activities including dining, games, movies, and tours increase the chances of close contact between passengers and crew [Fernandes 2014]. In addition, travel includes frequent layovers at ports of call where new crew and travelers can board, allowing the introduction of new susceptible persons and possibly new infected persons as well [Millman 2015]. Also, the modern cruise ships accommodate many travelers, often carrying passengers aged over 65 years, some with medical comorbidities, with an increased risk for complications. The incubation period and the subsequent period of maximum infectivity of many infectious diseases fall within the average cruise period of 6 days [Quigley 2020].

The high number of traveling individuals, frequently in close proximity to others, increases the likelihood of transmitting infectious diseases. As in other closed or semi-closed settings, the onboard transmission of viruses and consequent development of an outbreak is facilitated by direct person-to-person contact or contact with contaminated surfaces. The risk of transmission of infections depends on proximity to an index case, passengers and crew movement, fomites, and contact among passengers or crew.

During February – March 2020, SARS-CoV-2 outbreaks associated with cruise ship voyages caused over 800 cases among passengers and crew [Moriarty 2020]. Consequently, from February 2020, several national and international organizations provided recommendations on deferral of ship travel worldwide, and guidance on managing COVID-19 cases and outbreaks onboard ships [WHO 2020a, WHO 2020b]. In March 2020, the Cruise Lines International Association announced a 30-day voluntary suspension of cruise operations in the United States. Also, the CDC issued a No Sail Order for cruise ships, suspending operation in U.S. waters; the order was renewed on 9 April 2020, taking effect on 15 April 2020 [Scuchat 2020]. About 13 months later in the pandemic, guidance was provided on the gradual and safe resumption of operations of cruise ships [ECDC 2021, EU Health Gateways 2021].

One of the most critical aspects of these uncertainties is the modes and circumstances of transmission of newly identified agents. Nonetheless, given the novelty of the SARS-CoV-2 infection, early cruise ship travel transmission models of spread are based on what is known of the dynamics of other respiratory viral infections, mainly those due to influenza. Consequently, research is ongoing worldwide to understand SARS-CoV-2 modes of transmission, complemented with rapid publications. As a result, there is a need to continuously and systematically conduct reviews of publicly available studies with the latest knowledge to inform recommendations using the most up-to-date information.

## Objectives

The objectives of the review are to provide a rapid summary and evaluation of relevant data on the transmission of SARS-CoV-2 aboard cruise ships, report important policy implications, and highlight research gaps which need to be addressed. This includes airborne, contact and droplet, fomite, and orofecal modes of transmission.

### Modes of transmission for SARS-CoV-2 [WHO 2021, WHO 2014]

- Respiratory droplets are >5-10 μm in diameter. Respiratory droplets that include the virus can reach the mouth, nose, or eyes of a susceptible person and can result in infection.
- Respiratory droplets <5μm in diameter are referred to as droplet nuclei or aerosols. Airborne transmission is the spread of an infectious agent caused by disseminating aerosols that remain infectious when suspended in the air over long distances and periods of time.
- Close or direct contact transmission occurs with an infected person who has respiratory symptoms.
- Respiratory secretions or droplets can contaminate surfaces and objects, creating fomites (contaminated surfaces).
- Orofecal transmission occurs where the virus in fecal particles can pass from one person to the mouth of another. The main causes include lack of adequate sanitation and poor hygiene practices. Fecal contamination of food is another form of orofecal transmission.

### Subgroups

If feasible, the studies on the transmission of SARS-CoV-2 associated with cruise ship travel will form a subgroup of a broader systematic review on transmission dynamics of COVID-19 [Jefferson 2020]. We will report the evidence from studies with the results of reverse transcriptase-polymerase chain reaction (RT-PCR) reported by cycle threshold [Public Health England 2020], time from symptom onset, genomic sequencing (GS), and live culture of SARS-CoV-2, where available. Evidence from studies comparing culture with other means of diagnosis is not usually mode of transmission-specific. Updates of the culture review will be carried out alongside and in parallel with modes of transmission study extraction for all modes.

## Methods

This protocol is developed based on a previous protocol for a series of systematic reviews on the evidence on transmission dynamics of COVID-19 [Jefferson 2020].

### Search Strategy

We will search the following electronic databases: LitCovid and the WHO COVID-19 from inception, and medRxiv and Google Scholar databases from January 2020. Searches will be updated approximately every month; if the volume of new studies is significant, searches will be performed every fortnight. Search terms will include Covid-19, SARS-CoV-2, transmission, and cruise ship appropriate synonyms. In addition, reference lists of relevant articles, including reviews, will be screened for additional relevant studies.

### Study inclusion and exclusion

We will include studies reporting onboard SARS-CoV-2 transmission from passengers and/or crew to passengers and/or crew. We will consider any potential transmission mode, including droplet, airborne, fomite, fecal-oral, or other. Studies can be observational, including case series, ecological, or prospective; or interventional, including randomized trials and clinical reports, outbreak reports, case-control studies, experimental studies, and non-predictive modeling.

We will include studies that assess factors influencing transmission, such as infectivity of the index case (pre-symptomatic, asymptomatic or symptomatic, with or without wearing masks), the susceptibility of passengers (previous SARS-CoV-2 infection or vaccination, with or without wearing masks), and effectiveness of exposure (proximity to the index case, duration of exposure, technical specifications of the ship, quality of ventilation). Studies incorporating models to describe observed data will be included. Studies reporting solely predictive modeling will be excluded.

If two or more papers present the same data, we will include only the most comprehensive (or more recent/higher quality) report.

### Data extraction

We will include publication details (authors, year, country); cruise characteristics (origin and destination of the ship, duration of the voyage, technical specifications of the ship, stops, and information on ventilation system); data on the index cases (number, age, gender, country of residence or nationality, location on the ship, if they did or did not wear masks, symptoms during the cruise, laboratory confirmation of diagnosis); study type and details on contact tracing (definition of contact, secondary cases demographic data, symptoms, laboratory confirmation, contact tracing strategy, methods used to identify contacts, methods used for contacting contacts, total number of contacts identified, the total number of successfully traced contacts, the location of contacts in relation to the index case, immunological status and if they did not did not wear masks); exposure of primary and secondary cases (before, during, and after the cruise); conclusion on disease transmission (the number of cases/number of contacted passengers, and crew excluding index cases), intervention, and funding source of the study.

We will extract study data into data extraction templates Table 1 and Appendix 1 (study characteristics), Table 2 (methodological quality of studies), and a table with a summary of the main findings. References will be included in alphabetical order as a web appendix that facilitates updating. We will follow PRISMA reporting guidelines.

Data extraction will be performed by one author and independently checked by a second author. In case of disagreement, a third author will arbitrate.

### Quality assessment

We will assess the quality of included studies based on a modified QUADAS-2 tool using five domains [Jefferson 2020] (see Appendix Table 2). For studies that generate the hypothesis of onboard COVID-19 transmission, we will assess the strength of evidence for each study depending on the methods used to investigate the SARS-CoV-2 transmission [Jefferson 2021].

For the *onboard transmission studies*, we will investigate the following aspects:

1. a clearly defined setting, with a description of the ship and voyage, location of index cases, and secondary cases;
2. demographic characteristics (including age and gender) and sampling procedures adequately described, with data on the day of the sampling procedure and data on symptoms (including onset day);
3. follow-up strategy and duration sufficient for the outcomes; we will consider as adequate a comprehensive follow-up strategy, for at least 14 days;
4. the transmission outcomes assessed adequately; the studies should report the following data on secondary cases: demographic data, clinical data (with the day of onset of symptoms), and paraclinical data (RT-PCR with Ct <25; GS; viral cultures) with the day of the testing procedure;
5. main biases that are threats to validity taken into consideration, follow-up of >80% of passengers and crew, and exclusion of alternative exposures.

For the *environmental studies*, we will investigate the following aspects:

1. Does the description of methods contain sufficient detail to enable replication of the study;
2. Is the sample source clearly described;
3. Are the analyses and reporting appropriate;
4. Is the bias dealt with; did the authors acknowledge the potential biases and, if yes, did they made any attempts to address them;
5. Are there any applicability concerns?

Quality assessment will be performed by one author and independently checked by a second author. Disagreements will be resolved through discussion. If a resolution cannot be achieved, a third author will arbitrate.

### Data synthesis and reporting

The outcome will consist of the onboard cruise ships’ transmission of SARS-CoV-2. We will provide a narrative summary of the data and report the outcomes, including quantitative estimates where feasible and relevant. We will report the detection of a live culture of SARS-CoV-2 when reported (see also subgroups), and data on GS if available. Where possible, compatible datasets may be pooled for meta-analysis. We may write to authors for clarification of data and also report research and policy implications.

### Continual data release

Summary descriptions of important, relevant research papers identified are outlined in the tracker and corresponding folders in an ongoing manner. As important new data accumulates, we will produce a report as an individual rapid review and aim to make all our work available by depositing the review findings on the Oxford Research Archive. Where possible, we will submit the review report to relevant scientific journals for publication.

## Data Availability

All data produced in the present work are contained in the manuscript

## Conflict of interest statements

TJ was in receipt of a Cochrane Methods Innovations Fund grant to develop guidance on the use of regulatory data in Cochrane reviews (2015-018). In 2014–2016, he was a member of three advisory boards for Boehringer Ingelheim. TJ was a member of an independent data monitoring committee for a Sanofi Pasteur clinical trial on an influenza vaccine. TJ is occasionally interviewed by market research companies about phase I or II pharmaceutical products for which he receives fees (current). TJ was a member of three advisory boards for Boehringer Ingelheim (2014-16). TJ was a member of an independent data monitoring committee for a Sanofi Pasteur clinical trial on an influenza vaccine (2015-2017). TJ is a relator in a False Claims Act lawsuit on behalf of the United States that involves sales of Tamiflu for pandemic stockpiling. If resolved in the United States favor, he would be entitled to a percentage of the recovery. TJ is coholder of a Laura and John Arnold Foundation grant for development of a RIAT support centre (2017-2020) and Jean Monnet Network Grant, 2017-2020 for The Jean Monnet Health Law and Policy Network. TJ is an unpaid collaborator to the project Beyond Transparency in Pharmaceutical Research and Regulation led by Dalhousie University and funded by the Canadian Institutes of Health Research (2018-2022). TJ consulted for Illumina LLC on next generation gene sequencing (2019-2020). TJ was the consultant scientific coordinator for the HTA Medical Technology programme of the Agenzia per i Servizi Sanitari Nazionali (AGENAS) of the Italian MoH (2007-2019). TJ is Director Medical Affairs for BC Solutions, a market access company for medical devices in Europe. TJ was funded by NIHR UK and the World Health Organization (WHO) to update Cochrane review A122, Physical Interventions to interrupt the spread of respiratory viruses. TJ is funded by Oxford University to carry out a living review on the transmission epidemiology of COVID-19. Since 2020, TJ receives fees for articles published by The Spectator and other media outlets. TJ is part of a review group carrying out Living rapid literature review on the modes of transmission of SARS-CoV-2 (WHO Registration 2020/1077093-0). He is a member of the WHO COVID-19 Infection Prevention and Control Research Working Group for which he receives no funds. TJ is funded to co-author rapid reviews on the impact of Covid restrictions by the Collateral Global Organisation. TJ’s competing interests are also online https://restoringtrials.org/competing-interests-tom-jefferson

CJH holds grant funding from the NIHR, the NIHR School of Primary Care Research, the NIHR BRC Oxford and the World Health Organization for a series of Living rapid review on the modes of transmission of SARs-CoV-2 reference WHO registration No2020/1077093. He has received financial remuneration from an asbestos case and given legal advice on mesh and hormone pregnancy tests cases. He has received expenses and fees for his media work including occasional payments from BBC Radio 4 Inside Health and The Spectator. He receives expenses for teaching EBM and is also paid for his GP work in NHS out of hours (contract Oxford Health NHS Foundation Trust). He has also received income from the publication of a series of toolkit books and for appraising treatment recommendations in non-NHS settings. He is Director of CEBM and is an NIHR Senior Investigator.

DE holds grant funding from the Canadian Institutes for Health Research and Li Ka Shing Institute of Virology relating to the development of Covid-19 vaccines as well as the Canadian Natural Science and Engineering Research Council concerning Covid-19 aerosol transmission. He is a recipient of World Health Organization and Province of Alberta funding which supports the provision of BSL3-based SARS-CoV-2 culture services to regional investigators. He also holds public and private sector contract funding relating to the development of poxvirus-based Covid-19 vaccines, SARS-CoV-2-inactivation technologies, and serum neutralization testing.

JMC holds grants from the Canadian Institutes for Health Research on acute and primary care preparedness for COVID-19 in Alberta, Canada and was the primary local Investigator for a Staphylococcus aureus vaccine study funded by Pfizer for which all funding was provided only to the University of Calgary. He is co-investigator on a WHO funded study using integrated human factors and ethnography approaches to identify and scale innovative IPC guidance implementation supports in primary care with a focus on low-resource settings and using drone aerial systems to deliver medical supplies and PPE to remote First Nations communities during the COVID-19 pandemic. He also received support from the Centers for Disease Control and Prevention (CDC) to attend an Infection Control Think Tank Meeting. He is a member and Chair of the WHO Infection Prevention and Control Research and Development Expert Group for COVID-19 and a member of the WHO Health Emergencies Programme (WHE) Ad-hoc COVID-19 IPC Guidance Development Group, both of which provide multidisciplinary advice to the WHO and for which no funding is received and from which no funding recommendations are made for any WHO contracts or grants. He is also a member of the Cochrane Acute Respiratory Infections Working Group.

JB is a major shareholder in the Trip Database search engine (www.tripdatabase.com) as well as being an employee. In relation to this work Trip has worked with a large number of organisations over the years, none have any links with this work. The main current projects are with AXA and Collateral Global. He worked on Living rapid literature review on the modes of transmission of SARS-CoV-2 (WHO Registration 2020/1077093-0) and is part of the review group carrying out a scoping review of systematic reviews and meta-analyses of interventions designed to improve vaccination uptake (WHO Registration 2021/1138353-0).

ECR was a member of the European Federation of Neurological Societies (EFNS) / European Academy of Neurology (EAN) Scientist Panel – Subcommittee of Infectious Diseases (2013-2017). Since 2021, she is a member of the International Parkinson and Movement Disorder Society (MDS) Multiple System Atrophy Study Group, the Mild Cognitive Impairment in Parkinson Disease Study Group, and the Infection Related Movement Disorders Study Group. She was an External Expert and sometimes Rapporteur for COST proposals (2013, 2016, 2017, 2018, 2019) for Neurology projects. She is a Scientific Officer for the Romanian National Council for Scientific Research.

AP holds grants from the NIHR School for Primary Care Research.

IJO and EAS have no interests to disclose.

## References

European Centre for Disease Prevention and Control (ECDC). European Maritime Safety Agency. Guidance on the gradual and safe resumption of operations of cruise ships in the European Union in relation to the COVID-19 pandemic. Revision 1, 12 May 2021. Available at: https://www.ecdc.europa.eu/sites/default/files/documents/COVID-CRUISE-GUIDANCE-revision-1-May-2021.pdf

EU Health Gateways. Interim advice for restarting river cruise ship operations after lifting restrictive measures in response to the COVID-19 pandemic. Version 1, June 2021. Available at: https://www.healthygateways.eu/Portals/0/plcdocs/EU_HEALTHY_GATEWAYS_COVID-19_RESTARTING_INLAND_CRUISES.PDF

Fernandes EG, de Souza PB, de Oliveira ME, Lima GD, Pellini AC, Ribeiro MC, Sato HK, Ribeiro AF, Yu AL. Influenza B outbreak on a cruise ship off the São Paulo Coast, Brazil. J Travel Med. 2014; 21(5):298–303. doi: 10.1111/jtm.12132.

Jefferson T, Plüddemann A, Spencer EA, Brassey J, Heneghan C. The evidence on transmission dynamics of COVID-19: protocol for a series of Systematic Reviews. 2020. Available at https://www.cebm.net/evidence-synthesis/transmission-dynamics-of-covid-19/

Jefferson T, Heneghan C, Spencer E, Brassey J, Pluddeman A, Onakpoya I, Evans D, Conly JA. Hierarchical Framework for Assessing Transmission Causality of Respiratory Viruses. Preprints 2021, 2021040633. DOI: 10.20944/preprints202104.0633.v1.

Millman AJ, Kornylo Duong K, Lafond K, Green NM, Lippold SA, Jhung MA. Influenza Outbreaks Among Passengers and Crew on Two Cruise Ships: A Recent Account of Preparedness and Response to an Ever-Present Challenge. J Travel Med. 2015;22(5):306–311. doi:10.1111/jtm.12215

Moriarty LF, Plucinski MM, Marston BJ, Kurbatova EV, Knust B, Murray EL, et al. Public Health Responses to COVID-19 Outbreaks on Cruise Ships - Worldwide, February-March 2020. MMWR Morb Mortal Wkly Rep. 2020; 69(12):347–352. doi: 10.15585/mmwr.mm6912e3.

Public Health England. Understanding cycle threshold (Ct) in SARS-CoV-2 RT-PCR. A guide for health protection teams. 2020. Available at: https://www.gov.uk/government/publications/cycle-threshold-ct-in-sars-cov-2-rt-pcr

Quigley, A., Nguyen, P., Stone, H., Lim, S., MacIntyre, C. Cruise Ship Travel and the Spread of COVID-19 – Australia as a Case Study. International Journal of Travel Medicine and Global Health, 2020; 9(1): 10–18. doi: 10.34172/ijtmgh.2021.03

Schuchat A; CDC COVID-19 Response Team. Public Health Response to the Initiation and Spread of Pandemic COVID-19 in the United States, February 24-April 21, 2020. MMWR Morb Mortal Wkly Rep. 2020; 69(18):551–556. doi: 10.15585/mmwr.mm6918e2.

World Health Organization. Infection Prevention and Control of Epidemic-and Pandemic-prone Acute Respiratory Infections in Health Care. Geneva 2014 (available at https://apps.who.int/iris/bitstream/handle/10665/112656/9789241507134_eng.pdf;jsessionid=41AA684FB64571CE8D8A453C4F2B2096?sequence=1).

World Health Organization. Infection Prevention and Control of Epidemic-and Pandemic-prone Acute Respiratory Infections in Health Care. Geneva; 2014 (available at https://apps.who.int/iris/bitstream/handle/10665/112656/9789241507134_eng.pdf;jsessionid=41AA684FB64571CE8D8A453C4F2B2096?sequence=1).

World Health Organization. Roadmap to improve and ensure good indoor ventilation in the context of COVID-19. Geneva 2020. Available at Roadmap to improve and ensure good indoor ventilation in the context of COVID-19 (who.int). License CC BY-NC-SA 3.0 IGO

World Health Organization. Operational planning guidance to support country preparedness and response. Geneva: World Health Organization; 2020 https://www.who.int/publications/i/item/draft-operational-planning-guidance-for-un-country-teams

World Health Organization. Operational considerations for managing COVID-19 cases and outbreaks on board ships: interim guidance, 24 February 2020. Available at: https://apps.who.int/iris/handle/10665/331164 License: CC BY-NC-SA 3.0 IGO.

World Health Organization. Operational considerations for managing COVID-19 cases or outbreaks on board ships. Interim guidance 25 March 2020. Available at: https://www.who.int/publications/i/item/operational-considerations-for-managing-covid-19-cases-outbreak-on-board-ships

World Health Organization. Information Notice for IVD Users 2020/05. Nucleic acid testing (NAT) technologies that use polymerase chain reaction (PCR) for detection of SARS-CoV-2. Available at: https://www.who.int/news/item/20-01-2021-who-information-notice-for-ivd-users-2020-05

