## Supplementary material for "Transmission of SARS-CoV-2 associated with cruise ship travel: protocol for a systematic review (Version 1)": Table 2

| **Study** | **Study type** | **Description of methods with sufficient detail to replicate** | **Sample sources clear** | **Analysis and reporting appropriate** | **Is bias dealt with** | **Applicability** | **Notes** |
| --- | --- | --- | --- | --- | --- | --- | --- |

Table 2. Quality assessment of included studies.

Environmental studies

Human studies

| **Study** | **Study type** | **Clearly defined setting** | **Demographic characteristics / sampling procedures adequately described** | **Follow-up strategy and duration sufficient for the outcomes** | **The transmission outcomes assessed adequately** | **Main threats to validity taken into consideration?** | **Notes** |
| --- | --- | --- | --- | --- | --- | --- | --- |

1. *Setting* – ship and voyage description, location of index cases and secondary cases

2. *Demographic characteristics*: age, gender, sampling procedures (RT-PCR with Ct<25, GS, viral cultures), with day of procedure; symptoms (with onset day)

3. *Follow up* – comprehensive follow-up strategy, 14 days

4. *Transmission outcomes* – secondary cases: demographic data, clinical data (with day of onset of symptoms), paraclinical data (RT-PCR with Ct<25; GS, viral cultures), with day of procedure

5. *Bias* – follow up > 80%, alternative exposures excluded
